# Demographic characteristics and clinical features of patients presenting with different forms of cutaneous leishmaniasis, in Lay Gayint, Northern Ethiopia

**DOI:** 10.1101/2024.02.15.24302809

**Authors:** Endalew Yizengaw, Bizuayehu Gashaw, Mulat Yimer, Yegnasew Takele, Endalkachew Nibret, Gizachew Yismaw, Edward Cruz Cervera, Kefale Ejigu, Dessalegn Tamiru, Abaineh Munshea, Ingrid Müller, Richard Weller, James A. Cotton, Lloyd A. C. Chapman, Pascale Kropf

## Abstract

Cutaneous leishmaniasis (CL) is a neglected tropical disease caused by *Leishmania* parasites, that can cause long-term chronic disabilities. The clinical presentation of CL varies in both type and severity. CL presents as three main clinical forms: localised lesions (localised cutaneous leishmaniasis, LCL); mucocutaneous leishmaniasis (MCL) that affects the mucosa of the nose or the mouth; or as disseminated not ulcerating nodules (diffuse cutaneous leishmaniasis, DCL).

Here we recruited a cohort of CL patients in a newly established leishmaniasis treatment centre (LTC) in Lay Gayint, Northwest Ethiopia, and collected detailed demographic and clinical data.

The results of our study show that more males than females present to the LTC to seek diagnosis and treatment. 70.2% of CL patients presented with LCL and 20.8% with MCL. A small number of patients presented with DCL, recidivans CL (a rare form of CL where new lesions appear on the edges of CL scars) or with a combination of different clinical presentations. The duration of illness varied from 1 month to 180 months. Over a third of CL patients had additional suspected CL cases in their household. Despite the majority of CL patients having heard about CL, only a minority knew about its transmission or that it could be treated. Most CL patients lived in areas where environmental factors known to be associated with the transmission of CL were present.

This work highlights that CL is an important public health problem in Lay Gayint and emphasises the urgent need for more CL awareness campaigns, better health education and better disease management practices.

## INTRODUCTION

Cutaneous leishmaniasis (CL) is a neglected tropical disease caused by *Leishmania* parasites transmitted by sand fly vectors. It is present in Africa, the Americas, the Eastern Mediterranean, Europe, South-East Asia and the Western Pacific and is endemic in 90 countries [1]. In 2022, 205,662 new cases were reported, with the majority of cases reported from the Eastern Mediterranean [1]. However, due to the absence of CL awareness and reliable reporting systems in many countries, the real number of cases is likely to be much higher. For example, in Africa, out of 19 countries known to be endemic for CL, only 14 had reported cases in 2022 [1]. The disease can cause different clinical manifestations: localised CL (LCL), characterised by one or more ulcerating lesions; mucocutaneous CL (MCL), where the lesions affect the mucosa of the mouth and nose; diffuse CL (DCL), characterised by non-ulcerating nodules; and recidivans CL (RCL), where new lesions appear on the edges of CL scars. Because CL often leaves severe and permanent disfiguring scars, it is frequently associated with discrimination, stigma and substandard living conditions. The diagnosis of CL can be difficult, as it can cause lesions of similar appearance to other skin diseases such as leprosy, bacterial and fungal infections, and eczema [2]. Therefore, cases must be confirmed by identifying parasites in skin scrapings using microscopy or PCR. LCL is the most common form of the disease; it usually heals within one year. However, persistent LCL, MCL, DCL and RCL necessitate treatment and patients still experience frequent relapses [3]. The most commonly used treatments are antimonials; however two recent Cochrane reviews highlighted the low number of well-designed clinical trials that assessed the efficacy of antimonials, as well other treatments used to treat CL, and their long-term effects [4, 5].

Over 20 *Leishmania* species can cause CL: in the Old World, CL is mostly caused by *Leishmania (L.) tropica, L. major*, and *L. aethiopica*; and in the New World, by *L. braziliensis, L. mexicana and L. amazonensis* [6].

In Ethiopia, the majority of CL cases are caused by *L. aethiopica*; there have also been reports of CL caused by *L. tropica* and *L. major* [7]. CL transmission is thought to be mainly zoonotic, with hyraxes being the main reservoir host [8] and *Phebotomus* (P.) *longipes* and *P. pedifer* the most common vectors [8, 9]. Over 28 million individuals are at risk of CL, primarily in the highlands of Amhara, Oromia, Tigray and the Southern Nations, Nationalities and Peoples’ Region of Ethiopia [10]. While LCL is the most common form of CL in Ethiopia, MCL and DCL are relatively common, but reported percentages of the different clinical forms vary greatly between different studies [11].

Several studies have described the epidemiology of CL in Northern Ethiopia (summarised in [11]). In Tigray, a cross-sectional study performed from November 2011 to April 2012 showed a prevalence of CL of 14% [12]. At the Leishmaniasis Research and Treatment Centre, Gondar, Northern Amhara, a retrospective study showed that 1079 patients were diagnosed with CL over period of 10 years [13]. In these two studies, the different clinical presentations of CL were not identified. In the largest established CL treatment centre in eastern Amhara, in Boru Meda, 888 patients were diagnosed with CL from 2012 to 2018, the majority with LCL (89.2%), 6.9% with MCL and 3.9% with DCL [14]. However, 300km west of Boru Meda, in Lay Gayint where this study took place, no cases had been formally registered by the Amhara Regional Health Bureau until 2019; even though this area had been reported by health professionals to be endemic for CL. Following the establishment of a new Leishmaniasis Treatment Centre (LTC) in Lay Gayint hospital in 2019, and awareness campaigns, we published a preliminary study showing that large numbers of CL patients were identified in this area, with one of two clinical forms: 79.1% with LCL and 20.9% with MCL [15]. Yet, little is known about the demographic characteristics of patients presenting with the different forms of CL in this area. The aim of this study was to recruit a cohort of CL patients in Lay Gayint to provide detailed clinical description of the different forms of CL; as well as obtain detailed documentation of living habits, family history of CL, and environmental conditions and investigate how these factors may be associated with different CL presentations.

## MATERIALS AND METHODS

### Ethical approvals

This study was approved by the Research and Ethical Review Committee of the College of Science, Bahir Dar University (RCSVD 002/2011 EC), the National Research Ethics Review Committee of the Ministry of Science and Higher Education of Ethiopia (ref. No MoSHE/RD/ 14.1/10112/2020) and Imperial College Research Ethics Committee (ICREC 18IC4593). Informed written consent was obtained from each participant.

### Study area

This study was carried out in Nefas Mewcha Hospital, a primary hospital in Lay Gayint District, Northwest Ethiopia. Lay Gayint is found in the South Gondar administrative zone of the Amhara National Regional State (11° 50’ 59” N latitude and 38° 22’ 0” E longitude). The district of Lay Gayint has 9 health centres, 43 health posts, and 1 primary hospital, that are providing health care for an estimated population of 211,475 (projected from the latest official census in 2007). The district covers an area of about 1,522.4 km^2^, with a population density of 163.6 people/km^2^. The topography of the district is dominated by chains of mountains, hills, and valleys extending from the Tekeze river (1494m above sea level) to the Guna Mountain Summit (3991m above sea level). The annual mean minimum and maximum temperatures range from 8°C to 29°C; and the average annual rainfall of the district is 898.3mm.

### Cutaneous leishmaniasis patient recruitment

Following awareness campaigns [15], health extension workers identified individuals with potential CL lesions in their respective catchment areas and referred them to the LTC, in Nefas Mewcha Hospital. Some individuals with skin lesions also came after they heard about the LTC. All individuals were seen by a dermatologist who triaged them based on the clinical appearance of the lesions: a parasitological diagnosis was performed on potential CL patients and all other patients were referred for further tests.

### Diagnosis of CL

A parasitological diagnosis was used to confirm CL: a skin scraping was collected from the edge of the active lesion using a sterile scalpel. The scraping was smeared on a glass slide and stained with 10% Giemsa stain to identify and count the number of amastigotes by microscopy as described in [16]. The same grading system as that described in [16] was used to grade the number of amastigotes per slide. If the slide was negative, but the lesions had all the clinical features of CL as defined by The Guidelines for Diagnosis, Treatment and Prevention of Leishmaniasis in Ethiopia [2] (clinically suspicious lesion is defined as a skin nodule or ulcer with a raised edge appearing on someone who lives in an area known to be endemic for CL or visited such an area in the last 2 years), the patient was still considered to be a CL patient. Confirmed CL cases were treated with sodium stibogluconate i.m. (20 mg/kg/day) for 28 days, as described in the Guidelines for Diagnosis, Treatment and Prevention of leishmaniasis in Ethiopia.

### Collection of demographic and clinical data

A standardised interviewer-administrated questionnaire was used to collect socio-demographic and clinical information. For CL patients < 18 years old, their parent or guardian was asked to answer the questions. The duration of illness was defined as the time (in months) since the first lesion appeared. The number of lesions was counted by the interviewer and varied from 1 to >5 lesions. Body mass index (BMI) was measured by dividing body weight (kg) by the square of height (m).

### Knowledge about CL

To evaluate the knowledge of adult patients about CL, the following three questions were asked:

i. Had they heard about CL, locally named as “kuncher”?
ii. Did they know how the disease is transmitted, and if so, how?
iii. Did they know if the disease can be treated, and if so where?

### Statistical analysis

Data were evaluated for statistical differences as specified in the legend of each table and figure. Fisher’s exact test was used to test for associations between age or sex and CL type, and for a difference in the distribution of parasite gradings between adults and children. Mann-Whitney and Kruskal-Wallis tests were used to assess differences in illness duration and lesion number by CL type and Spearman’s rank correlation coefficient, *ρ*, was used to assess correlation between illness duration and lesion number. Differences were considered statistically significant at *p*<0.05. ∗=p<0.05, ∗∗=p<0.01, ∗∗∗=p<0.001 and ∗∗∗∗=p<0.0001. Unless otherwise stated, summary statistics given are medians followed by interquartile range (IQR) in square brackets.

## RESULTS

### Recruitment

In this study, we recruited 346 CL patients in a newly established Leishmaniasis Treatment Centre (LTC), in Nefas Mewcha Hospital, Lay Gayint. The recruitment took place from January 2019 to September 2022. 207 CL patients were adults and 139 were children. 72 adult patients were female with a median age of 35 [21.3-45.8] and 135 were male with a median age of 35 [21–52] (p=0.4542, Figure 1A). 57 child patients were female with a median age of 9 [7-12.5] and 82 were male with a median age of 11.5 [7–14] (p=0.3353) (Figure 1B). Adult patients came from 23, and children from 5, different districts (Figure 2 and Table 1). Most CL patients came from Lay Gayint (182 adult [87.9%] and 135 children [97.1%], Table 1).

**Figure 1:**
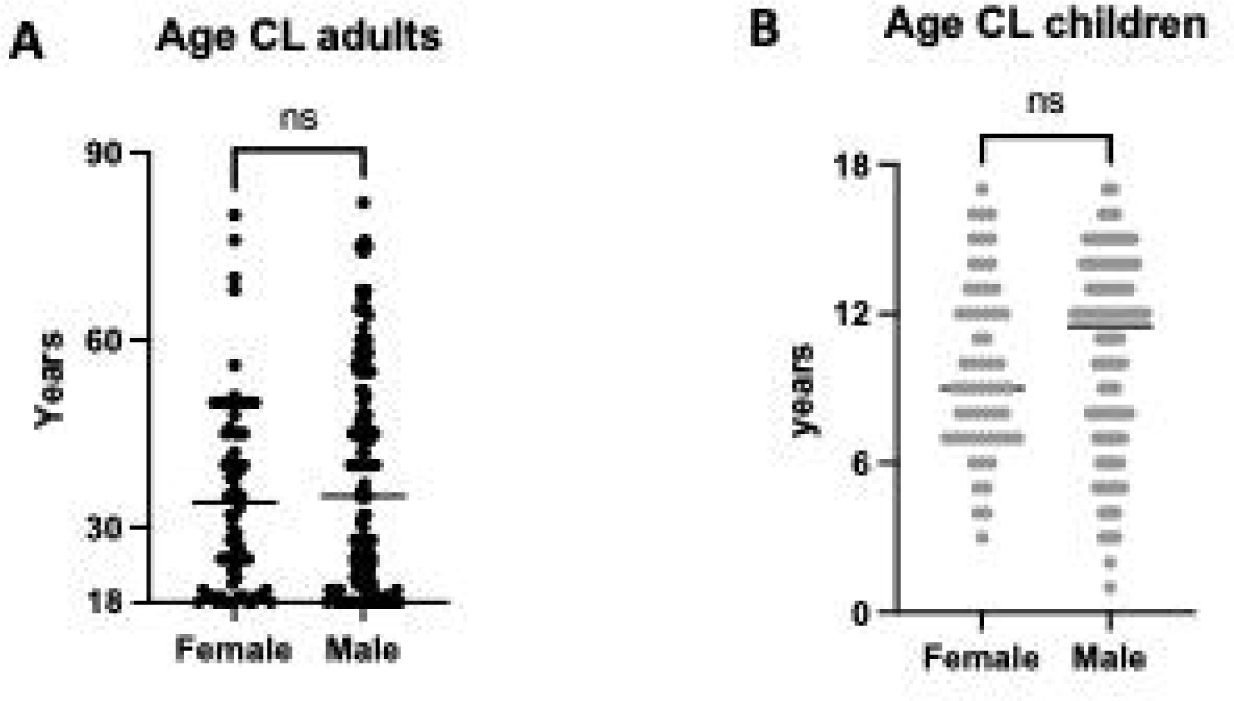
Ages of CL patients. (A) Ages of adult female (n=72) and male (n=135) CL patients. B. Ages of child female (n=57) and male (n=82) CL patients. Statistical differences were determined using a Mann-Whitney test. The straight line represents the median. ns=not significant.

**Figure 2:**
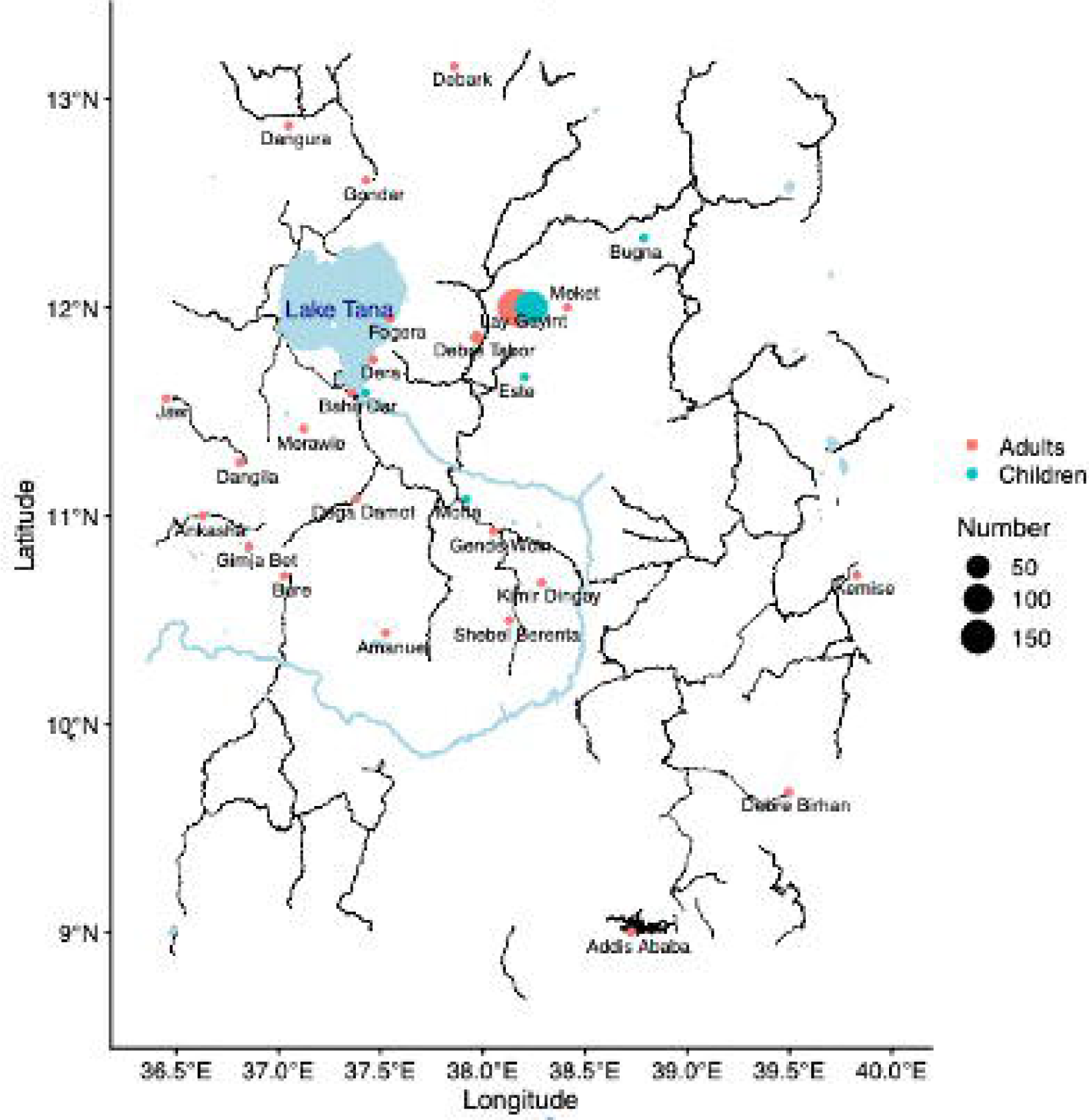
Map of the permanent places of residence of the recruited CL patients.

**Table 1:** permanent place of residence of CL patients.

| District/city | Adults | Children |
| --- | --- | --- |
| **Addis Ababa** | 1 | 0 |
| Amanuel | 1 | 0 |
| Ankasha | 1 | 0 |
| **Bahir Dar** | 1 | 1 |
| Bure | 1 | 0 |
| Bugna | 0 | 1 |
| Dangila | 1 | 0 |
| Dangura | 1 | 0 |
| Debark | 1 | 0 |
| Debre Birhan | 1 | 0 |
| Debre Tabor | 4 | 0 |
| Dega Damot | 1 | 0 |
| Dera | 1 | 0 |
| Este | 0 | 1 |
| Fogera | 1 | 0 |
| Gimja Bet | 1 | 0 |
| Gende Woin | 1 | 0 |
| Gondar | 1 | 0 |
| Jawi | 1 | 0 |
| Kemise | 1 | 0 |
| Kimir Dingay | 1 | 0 |
| Lay Gayint | 182 | 135 |
| Meket | 1 | 0 |
| Merawie | 1 | 0 |
| Motta | 0 | 1 |
| Shebel Berenta | 1 | 0 |

### Clinical forms of CL

CL patients presented with different clinical forms: localised CL (LCL), mucocutaneous CL (MCL), diffuse CL (DCL), and recidivans (RCL). LCL patients were further divided into two groups: those presenting with a well-defined contained lesion, with a distinct border around the lesion (contained LCL, C LCL) (Figure 3A) and those presenting with a lesion that did not have clear edges and was spreading (spreading LCL, S LCL) (Figure 3B). Some CL patients presented with multiple clinical forms of CL (multiple CL) (Table 2). Results presented in Table 2 show that the majority of adult and child CL patients presented with LCL (70% and 70.5%, respectively). There was no significant difference in the distribution of forms of CL between males and females, for either adults (p=0.2583) or children (p=0.3247, data not shown). Amongst adult and child LCL patients, there were more C LCL than S LCL (Table 3). 23.7% of adults and 16.5% of children presented with MCL (Table 2); 1.9% and 0.8% with DCL and 0.5% and 3.6% with recidivans CL. Eight adults and 12 children presented with multiple CL (Table 2).

**Figure 3:**
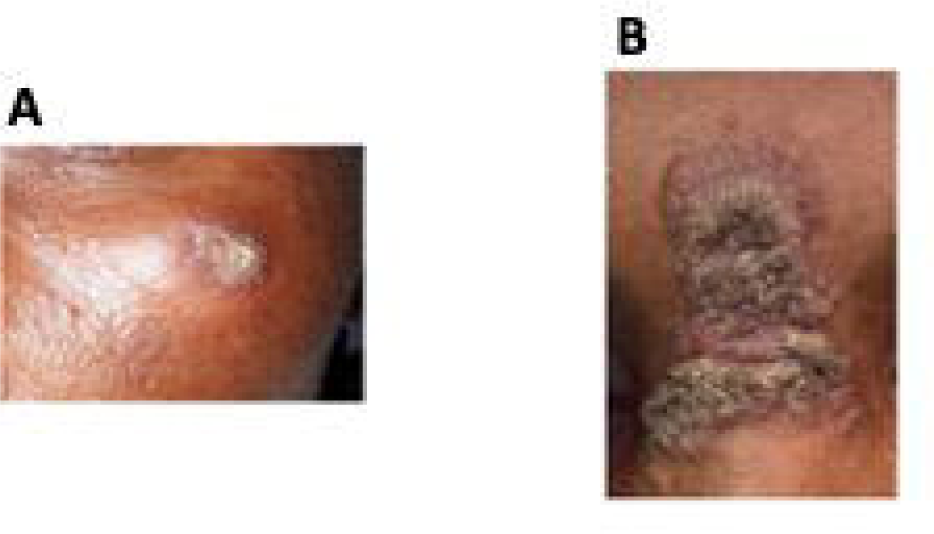
Examples of contained and spreading LCL lesions. **A.** Contained lesion, with a distinct border around the lesion (contained LCL, C LCL). **B.** Spreading lesion, without clear edges (spreading LCL, S LCL).

**Table 2:** Frequency of different clinical forms of CL.

| **Clinical presentations** | **Adults**<br>**n (%)** | **Children**<br>**n (%)** |
| --- | --- | --- |
| **LCL** | 145 (70) | 98 (70.5) |
| **MCL** | 49 (23.7) | 23 (16.5) |
| **DCL** | 4 (1.9) | 1 (0.8) |
| **RCL** | 1 (0.5) | 5 (3.6) |
| **Multiple CL\*** | 8 (3.9) | 12(8.6) |

\* 12 patients with MCL + C LCL, 4 with MCL + S LCL, 3 with S LCL + C LCL and 1 with DCL + MCL

**Table 3:** Number of contained and spreading LCL lesions.

|  |  |
| --- | --- |
|  | **Adults**<br>n (%) |
| **C LCL** | 105 (72.4) |
| **S LCL** | 40 (27.6) |
|  | **Children**<br>n (%) |
| **C LCL** | 72 (73.5) |
| **S LCL** | 26 (26.5) |

Amongst adults, the most represented age group of CL patients was 18-29 years. Amongst children, there was a similar number of CL patients in the 0-9 (n=62) and the 10-17 (n=77) age groups (Table 4). The most common form of CL was C LCL for all ages and sexes (Table 4). There was no association between age and form of CL for either adults (p=0.2237) or children (p=0.1475, data not shown).

**Table 4:** Frequency of different forms of CL by age group.

| **Children** | **C LCL**<br>**(n)** | **S LCL**<br>**(n)** | **MCL**<br>**(n)** | **DCL**<br>**(n)** | **RCL**<br>**(n)** | **Multiple**<br>**CL (n)** | **Total**<br>**(n)** |
| --- | --- | --- | --- | --- | --- | --- | --- |
| 0-9 | 27 | 16 | 10 | 0 | 4 | 5 | 62 |
| 10-17 | 45 | 10 | 13 | 1 | 1 | 7 | 77 |
| **Adults** | **C LCL**<br>**(n)** | **S LCL**<br>**(n)** | **MCL**<br>**(n)** | **DCL**<br>**(n)** | **RCL**<br>**(n)** | **Multiple**<br>**CL (n)** | **Total**<br>**(n)** |
| 18-29 | 45 | 23 | 19 | 3 | 0 | 2 | 92 |
| 30-39 | 20 | 5 | 2 | 0 | 1 | 0 | 28 |
| 40-49 | 16 | 4 | 15 | 1 | 0 | 3 | 39 |
| 50-59 | 17 | 4 | 7 | 0 | 0 | 0 | 28 |
| 60-69 | 9 | 4 | 2 | 0 | 0 | 1 | 16 |
| 70-79 | 3 | 0 | 3 | 0 | 0 | 1 | 7 |
| 80-89 | 1 | 0 | 1 | 0 | 0 | 0 | 2 |

### CL diagnosis (parasitological/clinical)

Parasitological diagnosis was used to confirm CL: 157 adults and 107 children tested positive by microscopy. The distribution of gradings of the number of amastigotes per slide is shown in Table 5. Most gradings were 1+ for both adults and children.

**Table 5:**
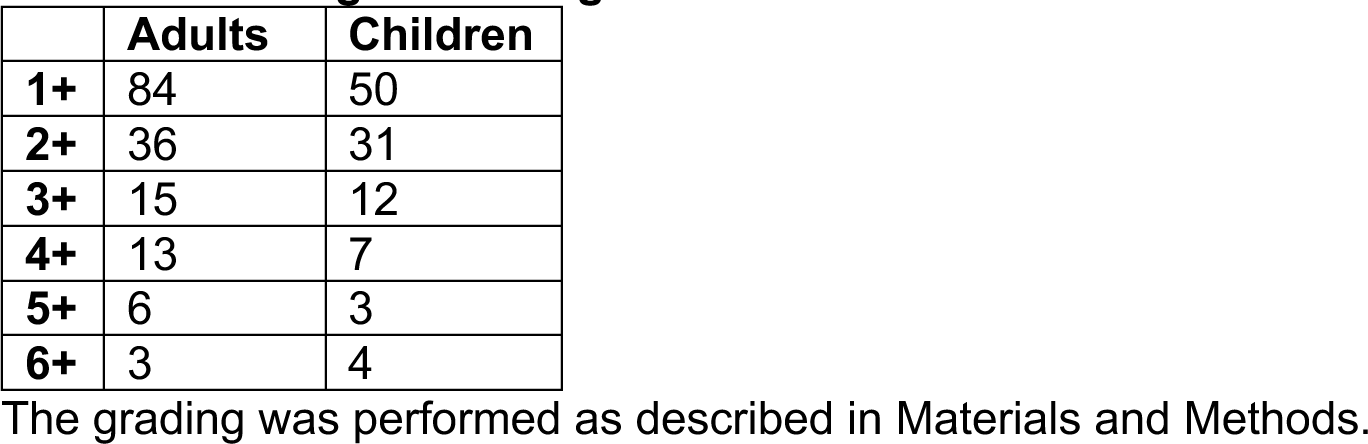
Grading of amastigotes.

There was no significant difference in the distribution of parasitological gradings between adults and children (p=0.8142). Fifty adults and 26 children tested negative but based on the appearance of their lesion(s) were considered to have CL by the dermatologist.

### Duration of illness

In adults, the duration of illness varied from 1 month to 180 months (Figure 4A and Table 6) and in children from 1-120 months (Figure 4B and Table 6). There were no significant differences in duration of illness between the different forms of CL in adults (p=0.0628, Figure 4A). In children, there was a significant difference in duration of illness between the different forms of CL in children (p=0.0301, Figure 4B), with the duration of illness in DCL patients being longer than in LCL (p=0.0315) and multiple CL (p=0.0217) patients. There was a similar trend in adult patients, however it was not significant (Figure 4A). There were no significant differences between the duration of illness in adult (p=0.1084) and child (p=0.8601) C LCL and S LCL patients (Figures 4C and D and Table 6).

**Figure 4:**
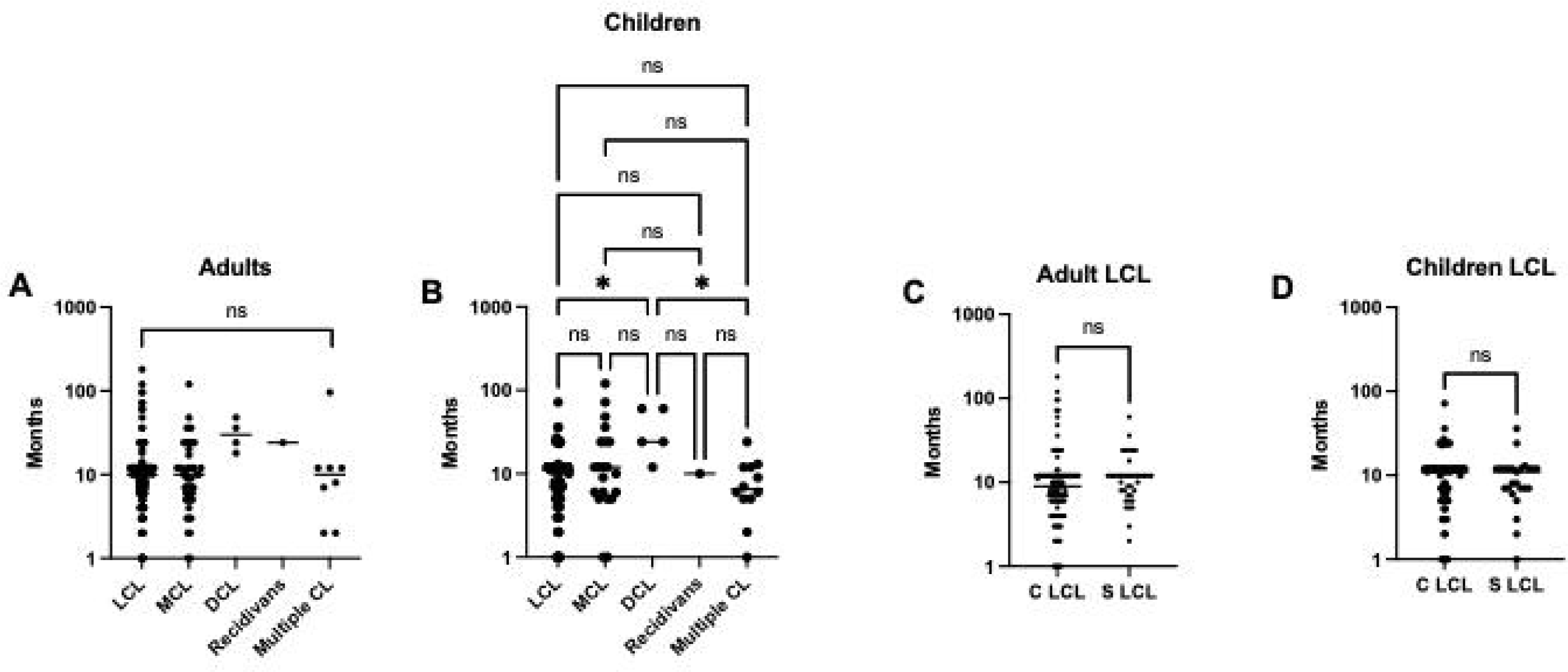
Duration of illness. **A.** Duration of illness in months for each clinical presentation in adult CL patients (LCL: n=144, MCL: n=49, DCL: n=4, recidivans: n=1, multiple CL: n=8). **B.** Duration of illness in months for each clinical presentation in child CL patients (LCL: n=94, MCL: n=21, DCL: n=5, recidivans: n=1, multiple CL: n=12). **C.** Duration of illness in months for each clinical presentation in adult S (n=105) and S (n=39) LCL patients. **D.** Duration of illness in months for each clinical presentation in adult S (n=69) and S (n=25) LCL patients. In Figures 4A and B, statistical differences between the five different clinical presentation of CL were measured by Kruskal-Wallis test and the multiple comparisons between each clinical presentation using Dunn’s multiple comparison test. In Figures 4C and D, statistical differences between S and C LCL were determined using a Mann-Whitney test. The straight line represents the median. ns=not significant.

**Table 6:** Duration of illness (in months) by form of CL.

|  | **LCL** | **MCL** | **DCL** | **Recidivans** | **Multiple CL** | **p values\*** |
| --- | --- | --- | --- | --- | --- | --- |
| **Adult** | 10 [6-12] | 10 [5.5-18.5] | 30 [19.5-45] | 24 [24-24] | 10 [3.3-12] | 0.0628 |
| **Children** | 11 [5.8-12] | 12 [5.5-24] | 24 [18-60] | 10 [10-10] | 6.5 [5-12] | **0.0301** |
|  | **C LCL** | **S LCL** | **p values#** |  |  |  |
| **Adult** | 9 [6-12] | 12 [7-18] | 0.1084 |  |  |  |
| Children | 11 [5-12] | 11 [7-12] | 0.8601 |  |  |  |

The durations of illness in adult CL patients were recorded for 144 LCL, 49 MCL, 4 DCL 1 recidivans CL and 8 multiple CL. For child CL patients, it was recorded for 94 LCL, 21 MCL, 5 DCL, 1 recidivans CL and 12 multiple CL. For the comparison between C LCL and S LCL, the durations of illness were recorded for 105 C LCL and 39 S LCL in adults and for 69 C LCL and 25 S LCL in children.
\*Statistical difference measured by Kruskal-Wallis

### Number of lesions

In adult and child patients, the numbers of lesions varied from 1 to >5 (Figure 5), and the majority (60.4%) of CL patients presented with one lesion. The highest numbers of lesions were identified in patients with DCL (Figure 5, Table 7).

**Figure 5:**
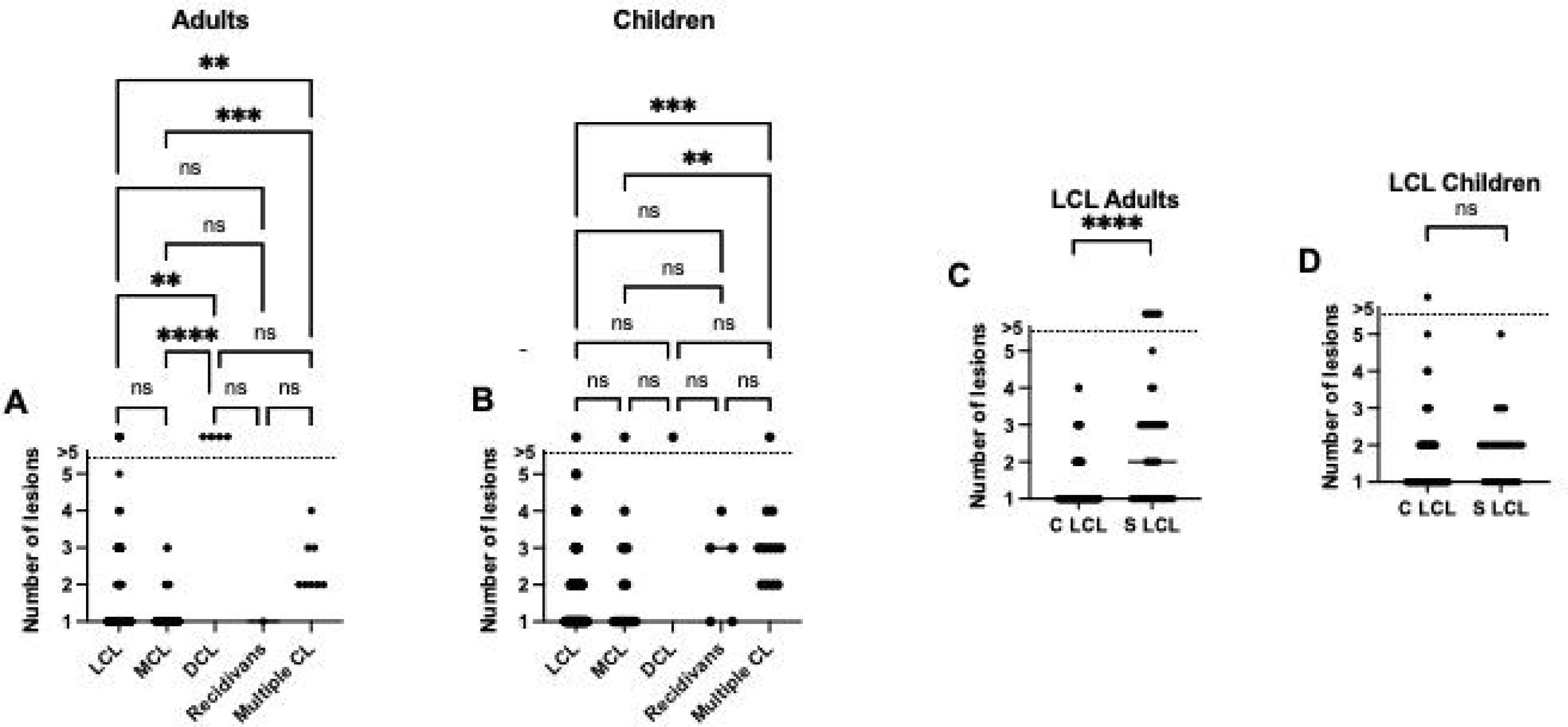
Number of lesions. **A.** number of lesions for each clinical presentation in adult CL patients (LCL: n=144, MCL: n=49, DCL: n=4, recidivans: n=1, multiple CL: n=8). **B.** Duration of illness in months for each clinical presentation in child CL patients (LCL: n=94, MCL: n=21, DCL: n=5, recidivans: n=1, multiple CL: n=12). **C.** Duration of illness in months for each clinical presentation in adult S (n=105) and S (n=39) LCL patients. **D.** Duration of illness in months for each clinical presentation in adult S (n=69) and S (n=25) LCL patients. In Figures 5A and B, statistical differences between the five different clinical presentations of CL were measured by Kruskal-Wallis test and the multiple comparison between each clinical presentation using Dunn’s multiple comparison test. In Figures 5C and D, statistical differences between S and C LCL were determined using a Mann-Whitney test. The straight line represents the median. ns=not significant.

**Table 7:** Number of lesions by CL type.

|  | **LCL** | **MCL** | **DCL** | **Recidivans** | **Multiple CL** | **p values\*** |
| --- | --- | --- | --- | --- | --- | --- |
| **Adult** | 1 [1-2] | 1 [1-1] | 6 [6-6] | 1 [1-1] | 2 [2-3] | <0.0001 |
| **Children** | 1 [1-1] | 1 [1-3] | 6 [6-6] | 3 [3-3.5] | 3 [2-3.8] | 0.0002 |
|  | **C LCL** | **S LCL** | **p values#** |  |  |  |
| **Adult** | 1 [1-1] | 1 [2-3] | <0.0001 |  |  |  |
| **Children** | 1 [1-2] | 1 [2-2] | 0.1006 |  |  |  |

The numbers of lesions in adult CL patients were recorded for 144 LCL, 49 MCL, 4 DCL 1 recidivans CL and 8 multiple CL. For child CL patients, it was recorded for 98 LCL, 23 MCL, 1 DCL, 5 recidivans CL and 12 multiple CL. For the comparison between C LCL and S LCL, the numbers of lesions were recorded for 104 C LCL and 40 S LCL in adults and for 72 C LCL and 26 S LCL in children.
\*Statistical difference measured by Kruskal-Wallis

There was a significant difference in the numbers of lesions between the different CL presentations in adults (Figure 5A, Table 7), with the numbers of lesions in LCL patients being lower than in DCL (p=0.0012) and multiple CL (p=0.0077) patients and the numbers of lesions in patients with MCL being lower than in DCL (p<0.0001) and multiple CL (p=0.0003) patients.

In children, there was also a significant difference in the numbers of lesions between the different CL presentations (Figure 5B, Table 7), with the numbers of lesions in patients with multiple CL being higher than in LCL (p=0.0003) and MCL (p=0.0046) patients.

The numbers of lesions were significantly higher in S LCL than C LCL in adults (Figure 5C, p<0.0001), but not in children (Figure 5D, p=0.1006).

There were weak positive correlations between the number of lesions and the duration of illness in adults and in children (*ρ*=0.246, p=0.0004 and *ρ*=0.241, p=0.0051, Table 8). When stratified according to the different clinical presentations, there was a weak positive correlation for adult patients with S LCL (*ρ*=0.382, p=0.0165) and for child patients with C LCL and MCL (*ρ*=0.266, p=0.0269 and *ρ*=0.640, p=0.0018) (Table 8).

**Table 8:** Correlations between the number of lesions and the duration of illness.

|  | **Rank correlation coefficient** | **p value<sup>‡</sup>** |
| --- | --- | --- |
| **Adults** |  |  |
| **CL (n=206)** | 0.246 | 0.0004 |
| **C LCL (n=105)** | 0.144 | 0.1460 |
| **S LCL (n=39)** | 0.382 | 0.0165 |
| **MCL (n=49)** | 0.162 | 0.2665 |
| **Children** |  |  |
| **CL (n=133)** | 0.241 | 0.0051 |
| **C LCL (n=69)** | 0.266 | 0.0269 |
| **S LCL (n=25)** | 0.010 | 0.9619 |
| **MCL (n=21)** | 0.640 | 0.0018 |

<sup>‡</sup>Spearman test

### Location of lesions

The majority of lesions were located on the face, with cheek and nose being the most affected areas (Table 9). A small number of patients had lesions on their ear, hand, thigh, shoulder or neck, or in multiple locations (Table 9).

**Table 9:**
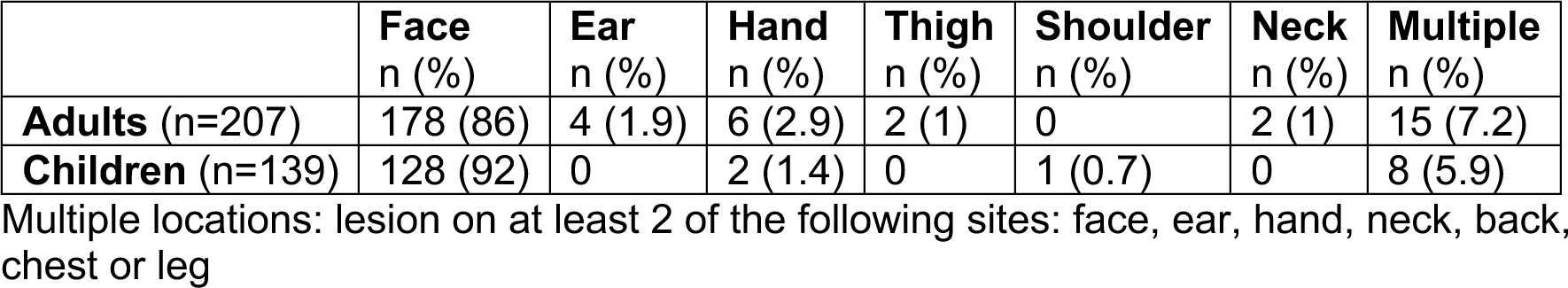
Location of lesions.

|  | **Face**<br>n (%) | **Ear**<br>n (%) | **Hand**<br>n (%) | **Thigh**<br>n (%) | **Shoulder**<br>n (%) | **Neck**<br>n (%) | **Multiple**<br>n (%) |
| --- | --- | --- | --- | --- | --- | --- | --- |
| **Adults** (n=207) | 178 (86) | 4 (1.9) | 6 (2.9) | 2 (1) | 0 | 2 (1) | 15 (7.2) |
| **Children** (n=139) | 128 (92) | 0 | 2 (1.4) | 0 | 1 (0.7) | 0 | 8 (5.9) |

Multiple locations: lesion on at least 2 of the following sites: face, ear, hand, neck, back, chest or leg

### CL patients’ occupations and education

There were four main occupations in adult CL patients (Table 10): farmer, government employee, student and merchant. The majority of patients were farmers (72%), followed by students (20.3%), government employees (5.8%) and merchants (1.9%).

**Table 10:** Occupation/education of adult CL patients.

| **Occupation**<br>**Education** | **Farmers**<br>**n (%)** | **Government employees**<br>**n (%)** | **Merchants**<br>**n (%)** | **Students**<br>**n (%)** |
| --- | --- | --- | --- | --- |
| **Total** | 149 (72) | 12 (5.8) | 4 (1.9) | 42 (20.3) |
| Illiterate | 87 (58.4) | NA | NA | NA |
| Can write and read | 16 (10.7) | NA | NA | 2 (4.8)* |
| Primary school | 41 (27.5) | NA | 3 (75) | 23 (54.8) |
| Secondary school | 2 (1.4) | 1 (8.3) | NA | 8 (19) |
| College and above | 3 (2) | 11 (91.7) | 1 (25) | 9 (21.4) |

\*Students attending Orthodox Church schools

The levels of education were also assessed and as shown in Table 10, the majority of CL patients were illiterate (58.4%). Students recruited in this study were studying at primary, secondary school, or college and above. Most children were in primary or secondary school (Table 11).

**Table 11:**
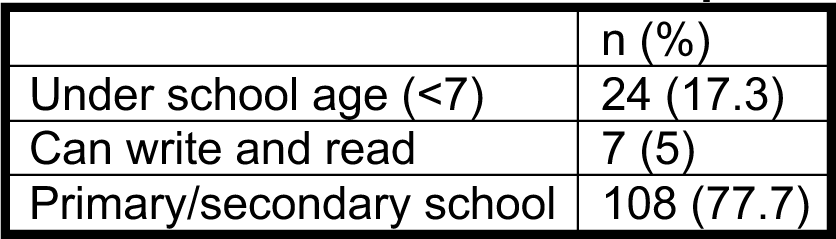
Education of child CL patients.

|  | n (%) |
| --- | --- |
| Under school age (<7) | 24 (17.3) |
| Can write and read | 7 (5) |
| Primary/secondary school | 108 (77.7) |

### BMI

The median BMIs for females (20.6 [18.5-22.5]) and males (21.0 [19.5-22.9]) were similar (p=0.0936, Figure 6A). As shown in Figure 6B, there was no statistically significant difference in BMI between the different clinical forms (p=0.1963).

**Figure 6:**
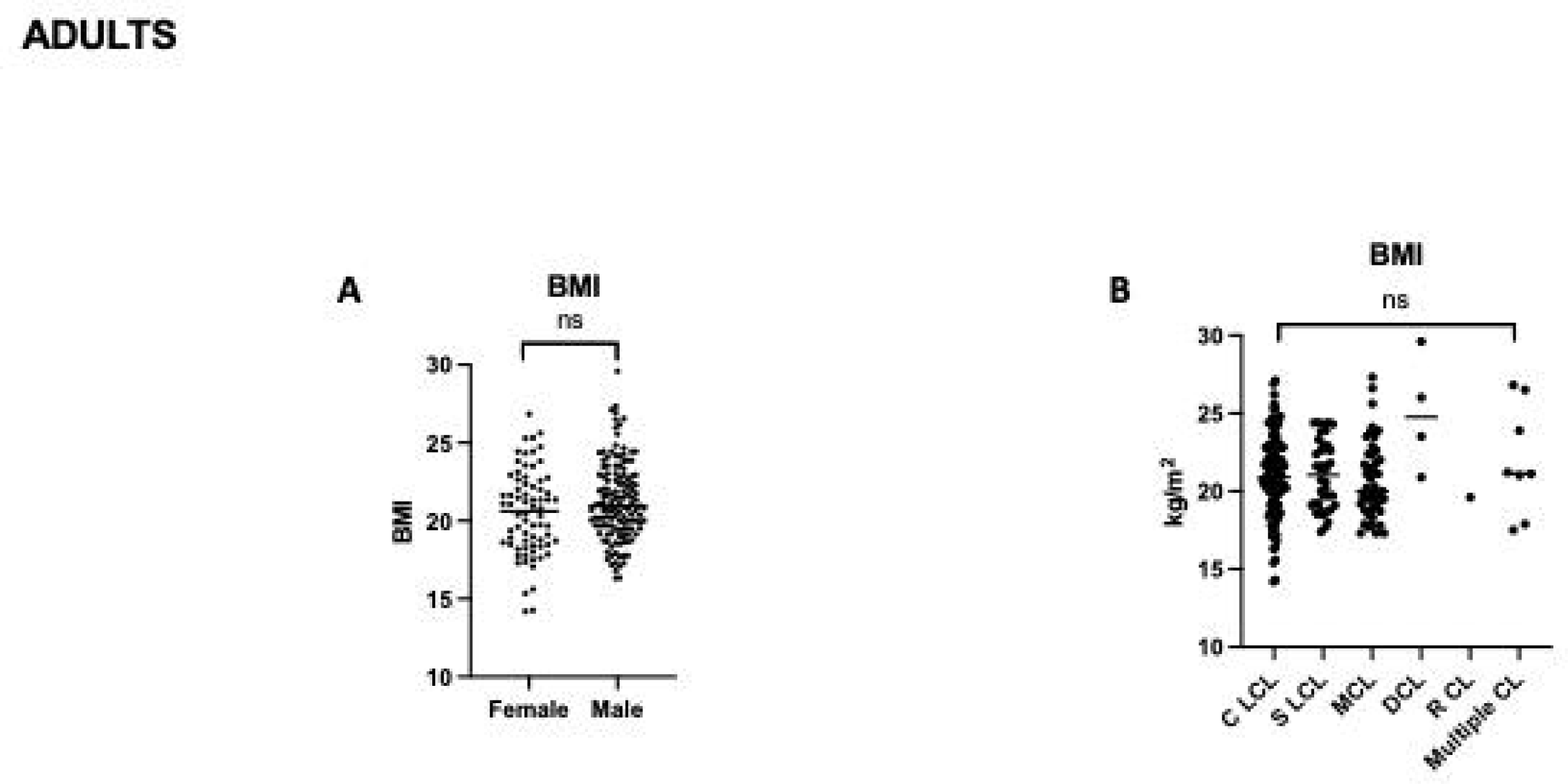
BMI per sex and clinical presentations. **A.** The BMI of male (n=72) and female (n=135) CL patients and **B.** the BMI of CL patients presenting with different clinical presentations (C LCL, n=105; S LCL, n=40; MCL, n=49; DCL, n=4; RCL, n=1; multiple CL, n=8) was determined as described in Materials and Methods. Statistical differences between the BMI in male and female (**A**) was determined using a Mann-Whitney test and between the different clinical presentations of CL (**B**) using a Kruskal-Wallis test. ns=not significant.

### History of CL in the household

207 adult CL patients were asked if someone else in the household was potentially presenting with CL lesions, as indicated by a health extension worker: 26 (12.6%) said that health extension workers had identified potential CL lesions (see Table 12 for details). For children, 52 guardians or parents were asked: 18 (34.6%) mentioned about a family member with suspected CL lesions (Table 12).

**Table 12:** Potential other CL cases in the households of diagnosed CL patients.

| **ADULTS** |  |  |  |  |  |  |
| --- | --- | --- | --- | --- | --- | --- |
| **Diagnosed CL patients** |  |  | **Potential CL cases** |  |  |  |
| **Age range of each patient** | **CL** | **Duration of illness** | **n** | **Age range of each patient** | **Duration of illness** | **Treatment** |
| 16-20 | S LCL | 12m | 2 | 36-40/21-25 | 12m | No |
| 16-20 | S LCL | 36m | 1 | 56-60 | 12m | Yes, unknown |
| 50-56 | C LCL | 7m | 1 | 56-60 | 5m | No |
| 36-40 | MCL | 20m | 1 | 11-15 | 3m | No |
| 31-35 | C LCL | 10m | 1 | 6-10 | 24m | Yes, traditional |
| 46-50 | S LCL | 12m | 1 | 26-30 | 12m | Yes, traditional |
| 21-25 | C LCL | 3m | 1 | 0-5 | 9m | No |
| 26-30 | C LCL | 4m | 1 | 26-30 | 2m | No |
| 35-40 | MCL | 12m | 1 | 11-15 | 24m | No |
| 16-20 | MCL | 7m | 1 | 26-30 | 6m | No |
| 56-60 | MCL | 120m | 3 | 16-20/6-10/6-10 | 12m | Yes, traditional |
| 56-60 | C LCL | 12m | 1 | 16-20 | 12m | Yes, traditional |
| 46-50 | MCL | 12m | 1 | 26-30 | 1m | Yes, unknown |
| 36-40 | Mixed | 2m | 1 | 31-35 | 2m | Yes, SSG |
| 61-65 | S LCL | 5m | 1 | 0-5 | 5m | No |
| 56-60 | C LCL | 4m | 1 | 50-56 | 7m | No |
| 41-45 | C LCL | 1m | 2 | 16-20/16-20 | 36m | No |
| 66-70 | S LCL | 7m | 1 | 0-5 | 6m | No |
| 46-50 | C LCL | 7m | 1 | 6-10 | 7m | No |
| 61-65 | C LCL | 10m | 1 | 50-56 | 12m | Yes, SSG |
| 18-20 | MCL | 2m | 1 | 36-40 | 12m | Yes, SSG |
| 26-30 | MCL | 2m | 1 | 26-30 | 4m | Yes, traditional |
| 71-75 | MCL | 4m | 2 | 36-40/50-56 | 12m | No |
| 26-30 | C LCL | 12m | 1 | 0-5 | 3m | No |
| 21-25 | C LCL | 24m | 1 | 16-20 | 12m | No |
| 31-35 | C LCL | 4m | 1 | 11-15 | 4m | No |
| **CHILDREN** |  |  |  |  |  |  |
| **Diagnosed CL patients** |  |  | **Potential CL patients** |  |  |  |
| **Age range of each patient** | **CL** | **Duration of illness** | **n** | **Age range of each patient** | **Duration of illness** | **Treatment** |
| 6-10 | MCL | 1m | 1 | 0-5 | 12m | No |
| 6-10 | C LCL | 7m | 1 | 46-50 | 7m | No |
| 11-15 | C LCL | 12m | 1 | 6-10 | 24m | No |
| 16-20 | MCL | 5m | 1 | 11-15 | 9m | No |
| 0-5 | S LCL | 12m | 1 | 6-10 | 1m | No |
| 11-15 | C LCL | 12m | 1 | 6-10 | 6m | No |
| 6-10 | S LCL | 3m | 1 | 11-15 | 12m | No |
| 6-10 | C LCL | 7m | 1 | 15 | 24m | No |
| 0-5 | MCL | 12m | 1 | 12 | 12m | Yes,<br>traditional |
| 6-10 | MCL | 6m | 1 | 14 | 12m | No |
| 11-15 | C LCL | 12m | 1 | 9 | 6m | No |
| 6-10 | S LCL | 7m | 1 | 5 | 7m | No |
| 11-15 | Mixed | 9m | 1 | 16 | 5m | No |
| 11-15 | C LCL | 24m | 1 | 54 | 12m | No |
| 6-10 | RCL | 60m | 1 | 14 | 24m | No |
| 11-15 | C LCL | 12m | 1 | 9 | 24m | No |
| 11-15 | C LCL | 24m | 1 | 2 | 48m | No |
| 0-5 | S LCL | 5m | 1 | 10 | 24m | Yes,<br>traditional |

Their age, the duration of illness and the treatment were also recorded (Tables 12). The majority had not received treatment yet.

### Knowledge about CL

Most CL patients (66.7%) had heard about CL, mainly from a member of their family or a friend (Table 13). A small percentage had heard about CL from a health facility or from school. Only 3.4% knew that CL is transmitted by an insect and 93.2% did not know that CL lesions can be treated (Table 13).

**Table 13:**
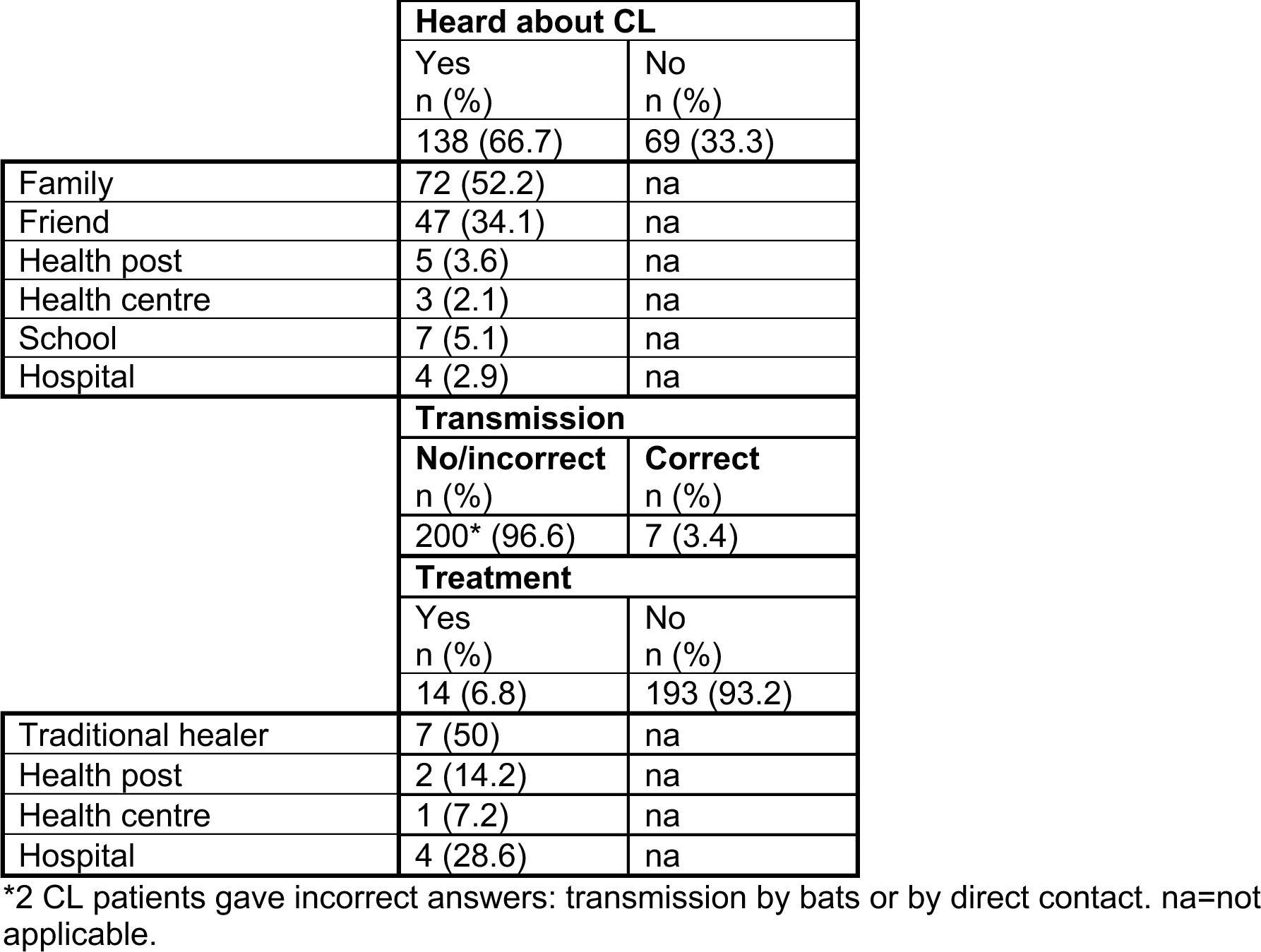
Knowledge about CL, transmission and treatment.

### Environmental factors

A total of 304 individuals (CL patients and parents or guardians of child patients) were asked about environmental factors known to be associated with the transmission of CL. The first question was about the presence of domestic animals such as dogs, goats, sheep, donkeys, cows, horses, cats, and/or chicken, as well as hyraxes, which are the main host reservoir for *L. aethiopica*. All CL patients had seen all these animals where they live or work, except for 23 CL patients, who had seen all the domestic animals but no hyraxes.

Next, they were asked if they live or work close to places where sand flies are known to breed, as well as places where hyraxes live such as caves, gorges, animal burrows, and or/ rock piles. All were present where the CL patients live or work.

Sand flies feed on the nectar, honeydew, and sap of certain trees, mainly *Acacia* and *Balanites* species. CL patients were asked to name the main trees present around the places where they live and work. Most CL patients (n=207) lived in the vicinity of *Acacia* and/or *Balanites*. Some other trees were also mentioned such as eucalyptus and juniper. 63 CL patients did not mention *Acacia* or *Balanites* but did mention tree species that are not associated with sand flies.

All houses where the CL patients lived had thatched grass wall, with cracks. All but one female and one male slept inside the house. 30 CL patients occasionally used bed nets, and none used an insect repellent.

## DISCUSSION

Our paper is the most detailed socio-demographic and epidemiological study to date of CL patients presenting in a recently established Leishmaniasis Treatment Centre (LTC), in Nefas Mewcha, Lay Gayint. CL in this area was poorly characterised. The District Health Office had described an outbreak of CL in Lay Gayint in 2009 and health professionals knew of CL cases from this part of Amhara. However, these had never been officially recorded by the Amhara Regional Health Bureau until 2019. We have recently published a study, describing the establishment of a new LTC in Nefas Mewcha in 2019 and reporting the retrospective data of a large number of CL patients presenting to this LTC [15]. This study highlighted that CL is a major health problem in this area.

In the current study, we recruited 207 adult and 139 child CL patients. This study took place over a 33-month period. The recruitment of patients was severely hampered by the COVID-19 pandemic and the civil war in Ethiopia. Due to these problematic recruitment conditions, patients were recruited as they came to the LTC. It is therefore not possible to infer the prevalence or incidence rate of active CL in adults or children in this area from these data.

As previously described there were more adult and child males than females presenting with CL [14, 15, 17, 18]. In our study this did not appear to be due to a difference in risk between males and females associated with different occupations as the majority of adult CL patients were farmers (who are at high risk of being bitten as they spend long periods of time working outside) and the proportion who were farmers among males and females was very similar (72.6% vs 70.9%, p=0.5020, data not shown). It is possible that men spend more time outside, as women oversee the cooking and might therefore spent more time indoors, and that this could explain why more males present with CL than females. However, it is also possible that men are more likely to seek diagnosis and treatment than women, at least in part because of the long and difficult journey to travel to the LTC. For some patients, the LTC was several days away from their villages.

As shown previously [15] and in agreement with previous studies in Ethiopia [11, 13, 19], the highest number of CL patients recruited were amongst children and young adults in the 18-29 age range. Due to the nature of recruitment, it is not possible to ascertain that this is representative of the age distribution of CL in this population. However, it has been shown that previous *Leishmania* infection can confer a high degree of protection against subsequent infection: leishmanization, where individuals are infected with virulent *L. major* parasites, has been shown to protect against subsequent natural infection in highly endemic areas [20]. It is therefore possible that in our cohort of CL patients, there are more younger individuals with active CL lesions, as older adults are likely to have been infected earlier in life and have become immune to reinfection.

LCL is the most common clinical presentation of CL worldwide [21], and Northern Ethiopia is no exception [11, 22]. Indeed, in our study in Lay Gayint, over 65% of adults and children presented with LCL. Our cohort of patients presenting with LCL was subdivided into two separate groups, those with a well-defined contained lesion (C LCL) that had a clear border and those presenting with a lesion that did not have distinct edges and was spreading (S LCL). The majority of LCL patients presented with C LCL, but almost a third with S LCL. It is tempting to speculate that nonhealing C LCL might become S LCL over time. However, we did not observe any difference in duration of illness. Since this was a cross sectional study and none of the patients were followed, we do not know the fate of C LCL lesions. A longitudinal follow-up from early lesions would allow for a better description of the clinical evolution of self-healing and spreading persistent lesions. MCL is usually quite rare, though the percentages of MCL reported by different studies vary greatly. In our cohort, the percentage of CL patients presenting with MCL was relatively high, 23.7% in adult patients and 16.5% in child patients. A study performed in Gondar, Northern Amhara, also showed a high percentage (42.7%) of patients presenting with MCL to the Leishmaniasis Research and Treatment Centre [22]. A small number of patients in our study also presented with different clinical presentations, with DCL or RCL. Since these forms of CL can be severely disfiguring, some of these patients hide their lesions and do not dare to seek diagnosis.

A considerable percentage of CL patients indicated that a health extension worker had identified other members of their family with potential CL lesions. This indicates that infection with *Leishmania* is not necessarily only associated with the occupation of the patients and could occur inside or close to the house [23, 24].

Most CL patients in our study had heard about CL, mainly from a member of their family, but very few from health facilities or even schools. Importantly, most did not know how it is transmitted and that it can be treated. This is in line with findings from other studies, where patients or individuals living in endemic areas know about the disease, but little about its transmission and treatments [25–28]. Better knowledge about how the disease is transmitted and the use of bed nets are likely to reduce CL transmission by sand flies. Indeed, it has been shown that systematic use of bed nets, in conjunction with indoor residual spraying, resulted in reduced incidence of CL in Mali [29]. The poor knowledge of the population studied here about treatment is likely to contribute to the long duration of illness and the severity of some of the lesions that were observed in this study. These often result in permanent disfiguration that can lead to social stigmatisation and a mental health burden [2]. Close follow-up of lesions to identify persistent nonhealing lesions that might benefit from early treatment is likely to both improve clinical outcomes [30] and reduce the risk of onward transmission.

The lack of knowledge about the extent of CL in this area, and in Ethiopia in general, is impeding effective control and prevention strategies. This work reinforces that CL is a major public health problem in Lay Gayint and emphasises the urgent need for more CL awareness campaigns, better health education and better disease management practices.

## Data Availability

All data produced in the present study are available upon reasonable request to the authors

## ACKNOWLEDGMENTS

The authors are thankful to staff of Nefas Mewcha Hospital for their enthusiastic collaboration during the data collection of this study.

This research is jointly funded by the UK Medical Research Council (MRC) and the Foreign Commonwealth and Development Office (FCDO) under the MRC/FCDO Concordat agreement (MR/R021600/1) (EY, BG, MY, JAC, LACC, PK).

## Notes

### Competing Interest Statement

The authors have declared no competing interest.

## REFERENCES

1. Ruiz-Postigo JA, Jain S, Madjou S, Virrey Agua JF, Maia-Elkhoury AN, Valadas S, et al. Global leishmaniasis surveillance, 2022: assessing trends over the past 10 years. Weekly epidemiological record. 2023;40(98):471–87.

2. Guidelines for diagnosis, treatment and prevention of leishmaniasis in Ethiopia, (2013). https://www.afrikadia.org/wp-content/uploads/2018/08/VL_Guidelines_Ethiopia_2013.pdf

3. Alvar J, Velez ID, Bern C, Herrero M, Desjeux P, Cano J, et al. Leishmaniasis Worldwide and Global Estimates of Its Incidence. PLoS ONE. 2012;7(5):e35671. Epub 2012/06/14. doi: 10.1371/journal.pone.0035671 PONE-D-11-24894 [pii]. PubMed PMID: 22693548; PubMed Central PMCID: PMC3365071.

4. Heras-Mosteiro J, Monge-Maillo B, Pinart M, Lopez Pereira P, Reveiz L, Garcia-Carrasco E, et al. Interventions for Old World cutaneous leishmaniasis. Cochrane Database Syst Rev. 2017;11(11):CD005067. Epub 20171117. doi: 10.1002/14651858.CD005067.pub4. PubMed PMID: 29149474; PubMed Central PMCID: PMCPMC6486265.

5. Pinart M, Rueda JR, Romero GA, Pinzon-Florez CE, Osorio-Arango K, Silveira Maia-Elkhoury AN, et al. Interventions for American cutaneous and mucocutaneous leishmaniasis. Cochrane Database Syst Rev. 2020;8(8):CD004834. Epub 20200827. doi: 10.1002/14651858.CD004834.pub3. PubMed PMID: 32853410; PubMed Central PMCID: PMCPMC8094931.

6. Burza S, Croft SL, Boelaert M. Leishmaniasis. Lancet. 2018;392(10151):951-70. Epub 20180817. doi: 10.1016/S0140-6736(18)31204-2. PubMed PMID: 30126638.

7. Hailu A, Di Muccio T, Abebe T, Hunegnaw M, Kager PA, Gramiccia M. Isolation of Leishmania tropica from an Ethiopian cutaneous leishmaniasis patient. Trans R Soc Trop Med Hyg. 2006;100(1):53–8. PubMed PMID: 16154167.

8. Ashford RW, Bray MA, Hutchinson MP, Bray RS. The epidemiology of cutaneous leishmaniasis in Ethiopia. Trans R Soc Trop Med Hyg. 1973;67(4):568–601. doi: 10.1016/0035-9203(73)90088-6. PubMed PMID: 4150462.

9. Lemma W, Erenso G, Gadisa E, Balkew M, Gebre-Michael T, Hailu A. A zoonotic focus of cutaneous leishmaniasis in Addis Ababa, Ethiopia. Parasit Vectors. 2009;2(1):60. Epub 2009/12/04. doi: 1756-3305-2-60 [pii] 10.1186/1756-3305-2-60. PubMed PMID: 19954530; PubMed Central PMCID: PMC2794267.

10. Seid A, Gadisa E, Tsegaw T, Abera A, Teshome A, Mulugeta A, et al. Risk map for cutaneous leishmaniasis in Ethiopia based on environmental factors as revealed by geographical information systems and statistics. Geospat Health. 2014;8(2):377–87. doi: 10.4081/gh.2014.27. PubMed PMID: 24893015.

11. van Henten S, Adriaensen W, Fikre H, Akuffo H, Diro E, Hailu A, et al. Cutaneous Leishmaniasis Due to Leishmania aethiopica. EClinicalMedicine. 2018;6:69–81. Epub 20190108. doi: 10.1016/j.eclinm.2018.12.009. PubMed PMID: 31193672; PubMed Central PMCID: PMCPMC6537575.

12. Bsrat A, Berhe N, Balkew M, Yohannes M, Teklu T, Gadisa E, et al. Epidemiological study of cutaneous leishmaniasis in Saesie Tsaeda-emba district, eastern Tigray, northern Ethiopia. Parasit Vectors. 2015;8:149. Epub 20150307. doi: 10.1186/s13071-015-0758-9. PubMed PMID: 25889827; PubMed Central PMCID: PMCPMC4359476.

13. Zeleke AJ, Derso A, Yeshanew A, Mohammed R, Fikre H. A Ten-Year Trend of Cutaneous Leishmaniasis at University of Gondar Hospital, Northwest Ethiopia: 2009-2018. J Parasitol Res. 2021;2021:8860056. Epub 20210309. doi: 10.1155/2021/8860056. PubMed PMID: 33777444; PubMed Central PMCID: PMCPMC7969101.

14. Eshetu B, Mamo H. Cutaneous leishmaniasis in north-central Ethiopia: trend, clinical forms, geographic distribution, and determinants. Trop Med Health. 2020;48:39. Epub 20200603. doi: 10.1186/s41182-020-00231-w. PubMed PMID: 32518497; PubMed Central PMCID: PMCPMC7271444.

15. Yizengaw E, Nibret E, Yismaw G, Gashaw B, Tamiru D, Munshea A, et al. Cutaneous leishmaniasis in a newly established treatment centre in the Lay Gayint district, Northwest Ethiopia. Skin Health Dis. 2023;3(4):e229. Epub 20230317. doi: 10.1002/ski2.229. PubMed PMID: 37538321; PubMed Central PMCID: PMCPMC10395643.

16. Chulay JD, Bryceson AD. Quantitation of amastigotes of Leishmania donovani in smears of splenic aspirates from patients with visceral leishmaniasis. Am J Trop Med Hyg. 1983;32(3):475–9. Epub 1983/05/01. PubMed PMID: 6859397.

17. Yohannes M, Abebe Z, Boelee E. Prevalence and environmental determinants of cutaneous leishmaniasis in rural communities in Tigray, northern Ethiopia. PLoS Negl Trop Dis. 2019;13(9):e0007722. Epub 20190926. doi: 10.1371/journal.pntd.0007722. PubMed PMID: 31557152; PubMed Central PMCID: PMCPMC6782111.

18. Haftom M, Petrucka P, Gemechu K, Nesro J, Amare E, Hailu T, et al. Prevalence and Risk Factors of Human Leishmaniasis in Ethiopia: A Systematic Review and Meta-Analysis. Infect Dis Ther. 2021;10(1):47–60. Epub 20201110. doi: 10.1007/s40121-020-00361-y. PubMed PMID: 33170497; PubMed Central PMCID: PMCPMC7652913.

19. Debash H, Ebrahim H, Bisetegn H. Epidemiological and clinical characteristics of cutaneous leishmaniasis among patients attending at Tefera Hailu Memorial Hospital, Sekota, Northeast Ethiopia: A five-year trend analysis (2016-2020). SAGE Open Med. 2022;10:20503121221129720. Epub 20221011. doi: 10.1177/20503121221129720. PubMed PMID: 36246535; PubMed Central PMCID: PMCPMC9558864.

20. Noazin S, Modabber F, Khamesipour A, Smith PG, Moulton LH, Nasseri K, et al. First generation leishmaniasis vaccines: a review of field efficacy trials. Vaccine. 2008;26(52):6759–67. Epub 20081023. doi: 10.1016/j.vaccine.2008.09.085. PubMed PMID: 18950671.

21. Scorza BM, Carvalho EM, Wilson ME. Cutaneous Manifestations of Human and Murine Leishmaniasis. Int J Mol Sci. 2017;18(6). Epub 20170618. doi: 10.3390/ijms18061296. PubMed PMID: 28629171; PubMed Central PMCID: PMCPMC5486117.

22. Fikre H, Mohammed R, Atinafu S, van Griensven J, Diro E. Clinical features and treatment response of cutaneous leishmaniasis in North-West Ethiopia. Trop Med Int Health. 2017;22(10):1293–301. Epub 20170813. doi: 10.1111/tmi.12928. PubMed PMID: 28712122.

23. Romero GA, Guerra MV, Paes MG, Macedo VO. Comparison of cutaneous leishmaniasis due to Leishmania (Viannia) braziliensis and L. (V.) guyanensis in Brazil: Clincial findings and diagnostic approach. CID. 2001;32:1304–12.

24. Jones TC, Johnson WD, Jr., Barretto AC, Lago E, Badaro R, Cerf B, et al. Epidemiology of American cutaneous leishmaniasis due to Leishmania braziliensis braziliensis. J Infect Dis. 1987;156(1):73–83. doi: 10.1093/infdis/156.1.73. PubMed PMID: 3598227.

25. Alharazi TH, Haouas N, Al-Mekhlafi HM. Knowledge and attitude towards cutaneous leishmaniasis among rural endemic communities in Shara’b district, Taiz, southwestern Yemen. BMC Infect Dis. 2021;21(1):269. Epub 20210317. doi: 10.1186/s12879-021-05965-4. PubMed PMID: 33731042; PubMed Central PMCID: PMCPMC7968254.

26. Ahmad S, Obaid MK, Taimur M, Shaheen H, Khan SN, Niaz S, et al. Knowledge, attitude, and practices towards cutaneous leishmaniasis in referral cases with cutaneous lesions: A cross-sectional survey in remote districts of southern Khyber Pakhtunkhwa, Pakistan. PLoS One. 2022;17(5):e0268801. Epub 20220526. doi: 10.1371/journal.pone.0268801. PubMed PMID: 35617283; PubMed Central PMCID: PMCPMC9135282.

27. Dires A, Kumar P, Gedamu S, Yimam W, Ademe S. Knowledge, attitude and prevention measures of students towards cutaneous leishmaniasis in Delanta district, Northeast Ethiopia. Parasite Epidemiol Control. 2022;17:e00241. Epub 20220125. doi: 10.1016/j.parepi.2022.e00241. PubMed PMID: 35146141; PubMed Central PMCID: PMCPMC8818578.

28. Alemayehu B, Kelbore AG, Alemayehu M, Adugna C, Bibo T, Megaze A, et al. Knowledge, attitude, and practice of the rural community about cutaneous leishmaniasis in Wolaita zone, southern Ethiopia. PLoS One. 2023;18(3):e0283582. Epub 20230328. doi: 10.1371/journal.pone.0283582. PubMed PMID: 36976758; PubMed Central PMCID: PMCPMC10047512.

29. Coulibaly CA, Traore B, Dicko A, Samake S, Sissoko I, Anderson JM, et al. Impact of insecticide-treated bednets and indoor residual spraying in controlling populations of Phlebotomus duboscqi, the vector of Leishmania major in Central Mali. Parasit Vectors. 2018;11(1):345. Epub 20180614. doi: 10.1186/s13071-018-2909-2. PubMed PMID: 29898753; PubMed Central PMCID: PMCPMC6000934.

30. WHO, Leishmaniasis. https://www.who.int/news-room/fact-sheets/detail/leishmaniasis.

